# Antigenic assessment of the H3N2 component of the 2019-2020 Northern Hemisphere influenza vaccine

**DOI:** 10.1101/2020.01.28.20019281

**Authors:** Sigrid Gouma, Madison Weirick, Scott E. Hensley

## Abstract

The 2019-2020 Northern Hemisphere influenza vaccine includes antigens from 3c3.A H3N2 viruses; however, over half of circulating H3N2 viruses belong to subclade 3c2.A1b. Here, we analyzed antibody responses elicited by the egg-adapted 3c3.A H3N2 vaccine strain in ferrets and humans. We found that this vaccine strain elicits antibodies that have reduced reactivity to a wild-type 3c3.A strain and very limited reactivity to 3c2.A strains, including the currently circulating 3c2.A1b strain.

---

H3N2 influenza viruses include many subclades with different genetic and antigenic properties^1^. Antigenically distinct 3c2.A and 3c3.A H3N2 viruses emerged during the 2014-2015 season^2^, which led to a major vaccine mismatch and low vaccine effectiveness^3,4^. 3c2.A H3N2 viruses have predominated since then, and antigens from these viruses have been included in vaccine formulations for the 2015-2016 to 2018-2019 seasons^1,5^. 3c2.A H3N2-based vaccines have been relatively ineffective^4^, partly because the hemagglutinins (HAs) of these viruses lose an antigenically important glycosylation site when propagated in eggs during the vaccine production process^6^. 3c3.A H3N2 viruses have circulated at low levels since the 2014-2015 season but unexpectedly dominated circulation in the United States during the late part of the 2018-2019 United States influenza season^1,7^. Due to the unexpected late circulation of 3c3.A H3N2 viruses, the World Health Organization (WHO) postponed their recommendation of 2019-2020 H3N2 vaccine strains during their vaccine strain selection meeting in February of 2019. A month later, the WHO recommended that antigens from 3c3.A H3N2 viruses should be included in the 2019-2020 formulation^7^. This was a controversial decision at the time since 3c2.A H3N2 viruses were circulating at high levels in most places in the world other than the United States^8^.

So far, very low levels of H3N2 viruses have circulated in the United States over the course of the 2019-2020 Northern Hemisphere influenza season^9^; however, H3N2 virus activity has been elevated in other parts of the world^10^. There has been significant diversity among currently circulating H3N2 viruses. For example, in Europe only 42% of sequenced H3N2 viruses from the 2019-2020 influenza season belong to the 3c3.A clade, while 58% belong to the 3c2.A clade^11^. Given the heterogeneity among circulating H3N2 viruses, we completed a series of experiments to assess the antigenic characteristics of the current 3c3.A H3N2 vaccine strain.

First, we analyzed antibodies from ferrets infected with either the egg-adapted A/Kansas/14/2017 3c3.A H3N2 vaccine strain or the wild-type A/Kansas/14/2017 3c3.A H3N2 virus. We tested antibodies raised against both wild-type and egg-adapted viruses because egg-adaptations can affect the antigenic properties of influenza vaccine strains^6,12^. The HAs of these two viruses differ by only 3 amino acids (**Figure 1A**). Ferrets infected with the wild-type 3c3.A strain produced antibodies that neutralized wild-type and egg-adapted 3c3.A viruses equivalently, whereas ferrets infected with the egg-adapted 3c3.A strain mounted antibody responses that efficiently neutralized egg-adapted 3c3.A virus and only moderately neutralized wild-type 3c3.A virus (**Figure 1B-C**). We used reverse-genetics to introduce each egg-adaptive HA substitution into wild-type 3c3.A viruses to determine which substitutions were responsible for the antigenic differences between the egg-adapted and wild-type viruses. The introduction of the egg-adapted G186V and D190N substitutions into the HA of wild-type 3c3.A viruses increased reactivity of antibodies elicited by the egg-adapted 3c3.A virus (**Figure 1C**). Both of these substitutions are located in antigenic site B (**Figure 1A**), indicating that 3c3.A viruses elicit an antibody response that is biased towards HA antigenic site B in ferrets.

**Figure 1.**
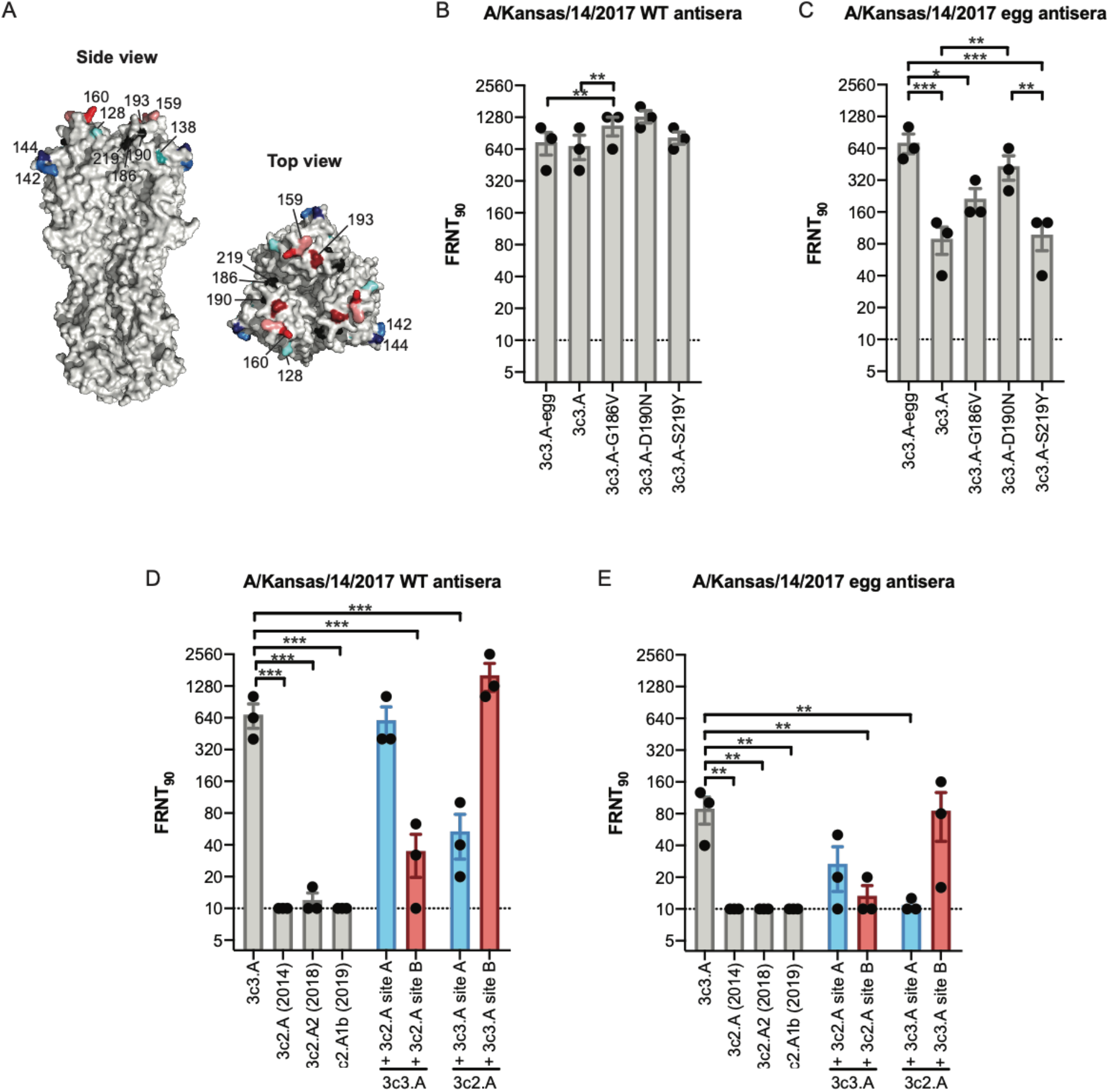
Antigenic mismatch of the H3N2 component of the 2019-2020 Northern Hemisphere influenza vaccine. (A) Crystal structure of the HA trimer of A/Victoria/361/2011 (PDB accession code 4O5I) with the amino acid substitutions that differ between egg-adapted and wild-type A/Kansas/14/2017 (positions 186, 190 and 219) shown in black and the amino acid substitutions that differ between wild-type A/Kansas/14/2017 and other 3c2.A H3N2 subclades in antigenic sites A (positions 128, 138, 142 and 144) and B (positions 159, 160 and 193) shown in blue and red, respectively. (B) Neutralizing antibody titers (FRNT_90_) to wild-type A/Kansas/14/2017 (3c3.A) and egg-adapted A/Kansas/14/2017 (3c3.A-egg) using wild-type A/Kansas/14/2017 antisera. (C) Neutralizing antibody titers (FRNT_90_) to wild-type A/Kansas/14/2017 (3c3.A) and egg-adapted A/Kansas/14/2017 (3c3.A-egg) using egg-adapted A/Kansas/14/2017 antisera. Data are presented as mean ± SEM. Significant differences are indicated. (D-E) Neutralizing antibody titers (FRNT_90_) to a panel of H3N2 viruses using wild-type A/Kansas/14/2017 and egg-adapted A/Kansas/14/2017 antisera. 3c3.A virus with 3c2.A site A possesses amino acid substitutions T128A, A138S, R142G and S144K. 3c3.A virus with 3c2.A site B possesses amino acid substitutions S159Y, K160T and S193F. 3c2.A virus with 3c3.A site A possesses amino acid substitutions A128T, S138A, G142R and K144S. 3c2.A virus with 3c3.A site B possesses amino acid substitutions Y159S, T160K and F193S. Data are presented as mean ± SEM. Significant differences relative to 3c3.A are indicated. * p<0.05, ** p<0.01, *** p<0.001.

HA antigenic site B of 3c3.A and 3c2.A H3N2 viruses are distinct since 3c2.A viruses possess a glycosylation site^6^ at HA residue 158 that is absent in 3c3.A viruses (**Figure 1A**). We therefore tested reactivity of the ferret antibodies to a panel of 3c2.A viruses, including a 3c2.A1b isolate that is representative of currently circulating 3c2.A H3N2 strains (**Figure 1D-E**). Antibodies from ferrets exposed to the wild-type or egg-adapted 3c3.A strain reacted poorly to all 3c2.A H3N2 viruses tested (**Figure 1D-E**). We used reverse genetics to engraft HA site A or B from 3c2.A viruses onto 3c3.A viruses and HA site A or B from 3c3.A viruses onto 3c2.A viruses. Antibodies from ferrets exposed to the wild-type or egg-adapted 3c3.A strain reacted more efficiently to viruses that possessed HA site B from 3c3.A viruses (**Figure 1D-E**), confirming that 3c3.A H3N2 elicits primarily an HA antigenic site B-focused response that fails to recognize 3c2.A HA.

Studies from our laboratory and others have demonstrated that ferrets and humans can have different anti-influenza virus antibody specificites, because most humans have extensive influenza virus immune histories^13^. We therefore analyzed serum antibodies collected from 62 humans before and after vaccination with an egg-based 2019-2020 Northern Hemipshere influenza vaccine. The median age of the participants in our study was 34 years (range 18-66 years) and 44 participants (71%) were female. Fifty-seven participants (92%) reported that they received at least one influenza vaccine during the last 2 years. Pre-vaccination antibody titers to wild-type 3c3.A virus, egg-adapted 3c3.A virus, and 3c2.A virus were low in most participants (**Figure 2A**). After vaccination, antibody titers to all viruses increased (**Figure 2A-B**), but titers against the egg-adapted 3c3.A strain were higher (GMT 209; 95% CI 156-281) compared to titers against the wild-type 3c3.A strain (GMT 78; 95% CI 58-104) and the 3c2.A1b strain (GMT 25; 95% CI 19-33) (**Figure 2A**). We found that 60 out of 62 participants (97%) had dectectable antibody titers to the 3c3.A H3N2 vaccine strain after vaccination, whereas only 35 out of 62 participants (56%) had detectable neutralizing antibody titers to 3c2.A1b H3N2 (**Figure 2A**). We also tested 3c3.A viruses that were engineered to possess either HA site A or site B from 3c2.A viruses (**Figure 2A-B**, blue and red respectively). Antibodies elicited by the vaccine reacted poorer to 3c3.A virus that possessed HA antigenic site B from 3c2.A viruses (**Figures 2A-B**), indicating that the vaccine mismatch between 3c3.A and 3c2.A viruses is primarily due to differences in HA antigenic site B.

**Figure 2.**
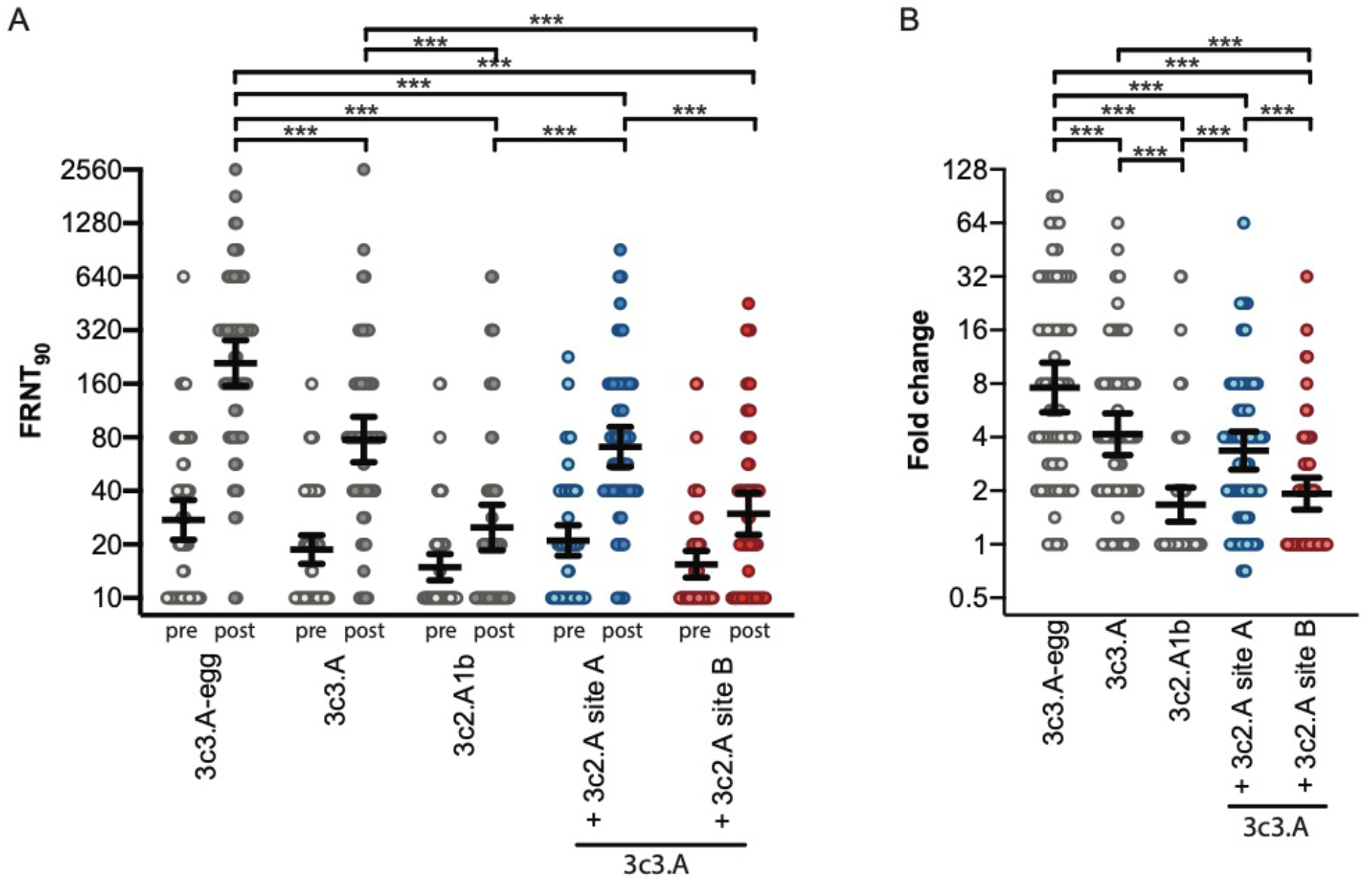
Neutralizing antibody titers (FRNT_90_) to a panel of H3N2 viruses in sera from adults (n=62) who received the 2019-2020 Northern Hemisphere influenza vaccine. A) Pre- and post-vaccination titers. B) Fold change of geometric mean neutralizing antibody titers (FRNT_90_) upon vaccination. 3c3.A virus with 3c2.A site A possesses amino acid substitutions T128A, A138S, R142G and S144K. 3c3.A virus with 3c2.A site B possesses amino acid substitutions S159Y, K160T and S193F. Significant differences are indicated. Thick horizontal lines show the geometric means and 95% confidence intervals. Post-vaccination titers were divided by pre-vaccination titers to calculate fold change. *** p<0.001.

Our within-season antigenic analyses indicate that the 3c3.A H3N2 component of the 2019- 2020 Northern Hemisphere influenza vaccine strain is antigenically mismatched to circulating 3c2.A H3N2 viruses. It will be important to continually monitor the frequencies of 3c3.A versus 3c2.A H3N2 viruses as the season progresses. It is not clear if one of these antigenically distinct H3N2 clades will predominate future influenza seasons, and therefore it will be difficult to make H3N2 component recommendations for future influenza vaccines. Ideally, antigens from both 3c3.A and 3c2.A H3N2 viruses will be included in future influenza vaccines; however, there are practical considerations that might not allow that to be immediately feasible. Since influenza vaccines protect against severe disease in years of antigenic mismatch^14^, the 2019-2020 Northern Hemisphere vaccine will likely offer some protection against both 3c3.A and 3c2.A H3N2 viruses.

## Methods

### Viruses

We used reverse-genetics^3^ to create viruses for this study. For ferret infections, we created 3c3.A viruses with the wild-type or egg-adapted A/Kansas/14/2017 HA (GISAID accession numbers EPI1146345 and EPI1444535). The egg-adapted HA possessed amino acid substitutions G186V, D190N, and S219Y relative to the wild-type HA (**Figure 1A**). All 3c3.A viruses were rescued using wild-type A/Kansas/14/2017 NA (GISAID accession number EPI1146344). Other viruses in our panel included HAs from A/Colorado/15/2014 (3c2.A, GISAID accession number EPI545672), A/North Carolina/28/2019 (3c2.A1b, GISAID accession number EPI1606044) or A/Pennsylvania/49/2018 (3c2.A2, GISAID accession number EPI1197412). NA from A/North Carolina/28/2019 (GISAID accession number EPI1606043) was used to create 3c2.A1b virus and NA from A/Colorado/15/2014 (3c2.A, GISAID accession number EPI545672) was used to create 3c2.A an 3c2.A2 viruses. Mutant viruses were created using either wild-type A/Kansas/14/2017 (3c3.A) or A/Colorado/15/2014 (3c2.A) HA as a backbone. 3c3.A virus with 3c2.A HA site A possesses HA amino acid substitutions A128T, S138A, G142R and K144S. 3c2.A virus with 3c3.A HA site A possesses HA amino acid substitutions T128A, A138S, R142G and S144K. 3c3.A virus with 3c2.A HA site B possesses HA amino acid substitutions S159Y, K160T and S193F. 3c2.A virus with 3c3.A HA site B possesses HA amino acid substitutions Y159S, T160K and F193S. All viruses were rescued using A/Puerto Rico/8/1934 internal genes to allow efficient viral growth in culture.

### Ferret sera

Ferrets were infected with 200,000 foci forming units (FFU) of virus with either wild-type A/Kansas/14/2017 (n=3) or egg-adapted A/Kansas/14/2017 (n=3) HA, and then bled 28 days later. Ferrets tested negative for influenza-specific antibodies in ELISA prior to infection. All animal experiments were completed under an Institutional Animal Care and Use Committee-approved protocol at Noble Life Sciences (Gaithersburg, MD).

### Human sera

Paired serum samples were collected from 62 participants at the University of Pennsylvania 1-12 days before and 26-33 days after vaccination in October 2019 with the 2019-2020 Northern Hemisphere influenza vaccine. All participants received either Fluarix (n=60) or Fluzone (n=2), which are both quadrivalent egg-based vaccines. Informed consent was obtained from all participants. The study was approved by the institutional review board of the University of Pennsylvania.

### Foci Reduction Neutralization Tests (FRNTs)

Serum samples were treated with receptor-destroying enzyme (Denka Seiken) followed by heat-inactivation at 55°C for 30 minutes prior to testing in FRNTs. FRNTs were performed as previously described^6^. Each ferret sample was tested in 3 independent experiments and each human sera was tested in 2 independent experiments. Geometric mean titers of the replicates were used for analysis.

### Statistical methods

Log_2_-transformed antibody titers measured in ferret and human sera were compared using RM one-way ANOVA corrected for multiple comparisons (Bonferroni method). Statistical analysis was performed using Prism version 8 (GraphPad Software) and R version 3.5.3 (R Foundation for Statistical Computing).

## Data Availability

All data is presented in the manuscript.

## Acknowledgments

This work was funded by institutional funds from the University of Pennsylvania and the National Institute of Allergy and Infectious Diseases at the National Institutes of Health (R01AI113047 (S.E.H.), R01AI108686 (S.E.H.), and HHSN272201400005C (S.E.H.). S.E.H. holds Investigators in the Pathogenesis of Infectious Disease Awards from the Burroughs Wellcome Fund. We thank the participants that took part in this study and the Penn Center for Human Phenomic Science unit for assisting in human blood draws.

## Author contributions

S.G. and S.E.H. designed the study. S.G. and M.W. completed experiments. S.G. and S.E.H. analyzed data and wrote the manuscript.

